# Lactoferrin levels in cerebrospinal fluid exhibits isoform-specific associations with Alzheimer’s disease

**DOI:** 10.1101/2025.09.24.25336349

**Authors:** Feiyang Zhao, Raquel Puerta, Yaxi Wang, Eva Beckett, Pablo Garcia Gonzalez, Sergi Valero, Adelina Orellana, Pilar Sanz, Tiffany F. Kautz, Jose E. Cavazos, Maria Victoria Fernandez, Amanda Cano, Sudha Seshadri, Merce Boada, Valentina R. Garbarino, Agustin Ruiz

## Abstract

**Background:** Pathological changes in Alzheimer’s disease (AD) begin years before the onset of clinical symptoms. Developing cost-effective and minimally invasive biomarkers for preclinical diagnosis remains a critical goal in the field. Lactoferrin, an iron-binding glycoprotein, has emerged as a promising candidate due to its multifunctional roles reported in previous studies. However, whether lactoferrin levels in biofluids are associated with established AD biomarkers, and whether it can serve as a reliable diagnostic indicator, remains controversial.

**Methods:** We analyzed SOMAscan proteomic data from 1,367 paired plasma and cerebrospinal fluid (CSF) samples from the ACE Alzheimer’s Center Barcelona cohort to evaluate lactoferrin levels. Associations between two lactoferrin-targeting SOMAmers and classical AD biomarkers, including total tau (t-tau), phosphorylated tau (p-tau181), and amyloid-beta 42 (Aβ42), were assessed. The age, sex, proteomic principal components were considered as covariates for sensitive analysis. The prognostic value of lactoferrin in predicting dementia progression was further evaluated using survival analysis.

**Findings:** Among the two lactoferrin-targeting SOMAmers, Seq.2780.35 (LTF2) showed a weak and exclusive association with CSF Aβ42 and syndromic status, whereas Seq.14755.4 (LTF1) was weakly associated with CSF p-tau and t-tau AD biomarker levels displayed expression-dependent stratification consistent with a ventricular-volume–related dilution effect rather than disease. Furthermore, lactoferrin levels were not significantly associated with the progression from mild cognitive impairment (MCI) to dementia.

**Interpretation:** Isoform-specific lactoferrin expression changes in cerebrospinal fluid (CSF), but not plasma, appears to have biological relevance and diagnostic biomarker potential for AD.

## Introduction

Alzheimer’s disease (AD) is the most common cause of dementia in the elderly, which is characterized by progressive cognitive decline, loss of neurons, amyloid-beta (Aβ) deposition, and tau pathology (1, 2). Combinations of clinical assessment, structural/functional neuroimaging, and evaluation of cerebrospinal fluid (CSF) based pathological markers, are considered well-established AD biomarkers and show a good diagnostic precision to detect AD progression (2, 3). Despite recent advancements in diagnostic tools, these CSF and imaging-based markers are either invasive or expensive, highlighting an urgent need for new diagnostic biomarkers to address these limitations. Furthermore, the underlying cause of neurodegenerative disease remains unclear. Even the widely accepted hypothesis, that Aβ aggregation leads to a pathological cascade, cannot alone fully explain the pathological initiation and progression of AD (3, 4). In the last decades, there has been growing recognition that immune system declines during aging and persistent inflammation induced by microbes contribute to Aβ deposition, progressive synaptic dysfunction, neuronal loss, and ultimately AD (5). In support of this hypothesis, infections of the central nervous system are known to be associated with AD-like pathology (6–10). Furthermore, oral and gut microorganisms are now known to play a role in the cascade of immune dysregulating events preceding AD (11, 12). The involvement of pathogen-targeting agents and viral infection–related markers in the brain such as amyloid aggregation, reinforces the potential association between AD and immunologic response of the central nervous system to infection(s) (12, 13). These pieces of evidence suggest an important role of both local and systemic inflammation towards AD pathogenesis.

As primary effector molecules of innate immunity, antimicrobial proteins and peptides may contribute significantly to AD development (14, 15), specifically Aβ pathology (16–19). Lactoferrin (LTF), an iron-binding glycoprotein involved in innate immunity, acting as part of the body’s first line of defense against bacterial, viral, fungal, inflammatory and carcinogenic compounds (20), has attracted attention in recent years for its potential role in neurodegeneration (21–23). Beyond exocrine secretion by epithelial cells in most mucosal tissues, it is also expressed by the cells of the innate immune system throughout the body, including peripheral neutrophil leukocytes and monocytes, and microglial within the brain (24, 25). In neutrophils, LTF is stored in the secondary granules and released into the blood or infected tissues at high concentrations during inflammation. LTFs expression and release occur not only in peripheral tissues but also in the central nervous system, where microglial cells can release LTF through degranulation, particularly under inflammatory conditions (25, 26).

In the last decade, several studies have reported altered LTF abundance in biological fluids of AD patients. For instance, through a cross-sectional investigation, Carro, et al. found that salivary LTF expression can discriminate AD patients from healthy controls more accurately than Aβ42 and total-tau (t-tau) in CSF (27). The same research group further revealed that salivary LTF can reliably distinguish prodromal AD and clinical AD groups from frontotemporal dementia (FTD) with high accuracy (28). Through a later study, they also demonstrated negative associations between salivary LTF and Aβ load, middle temporal cortex thickness, increased FDG uptake in posterior cingulate cortex, and poorer memory (29). Studies from this research group have consistently validated their previous findings, that salivary LTF may be a promising non-invasive and cost-effective AD biomarker. However, a subsequent, independent follow-up study in a mixed memory clinic cohort did not find an association between LTF levels in CSF or saliva and cognitive diagnoses or pathological AD markers (t-tau, p-tau, Aβ) (30). These conflicting findings from different research groups highlight the need for standardization across sample collection and assessment methods, and suggest that LTF may have a complex, context-dependent relationship with specific neurodegenerative disease pathology and progression—relationships which are not fully captured or detectable by current studies and evaluation methods. Disagreement between these studies may stem from several complex aspects of neurodegenerative disease pathology and the biology of LTF that is endogenously generated by a variety of cell types including: immune cell, glial cells, epithelial cells (25, 31). LTF not only exists in the neural system but also within the immune, digestive, circulatory and reproductive systems, so it is important to consider that circulating LTF levels are potentially modulated by multiple tissues, environmental exposures and potentially even some genetic factors.

To explore the potential involvement of LTF in AD and strengthen our understanding of LTFs role in AD, we examined data from a novel and independent cohort. We utilized existing proteomic data from the ACE Alzheimer Center Barcelona (Spain), which includes a large number of participants with paired CSF and plasma proteomic profiles measured by the high-throughput SOMAscan 7K platform. By leveraging this well-characterized dataset along with readily available detailed clinical phenotyping, we aimed to clarify the relationship between LTF and AD across CSF and plasma, and to assess whether LTF holds promise as a disease-associated or stage-specific biomarker in these two biofluids. This study provides an opportunity to re-evaluate the role of LTF in AD pathogenesis with improved statistical robustness and biological resolution.

## 2. Material and methods

### 2.1. Study Cohort

We used the ACE Memory Clinic in Barcelona, Spaine cohort of well characterized CSF and matched plasma samples for all analysis in this study. Participants who presented for cognitive evaluation and met criteria for lumbar puncture were recruited. On the same day, both biological samples were collected, along with clinical, cognitive, and imaging data. Follow-up assessments were conducted annually. The cohort consisted of individuals with MCI, dementia, or subjective cognitive concerns. Diagnoses were made by a multidisciplinary team based on a comprehensive neuropsychological evaluation. Progression to dementia was monitored over time to classify participants as either converters or stable. Converters are defined as subjects who converted to dementia, including AD, vascular dementia, mixed dementia (AD with cerebrovascular disease), frontotemporal dementia, or dementia with Lewy bodies over the study period (from the baseline assessment to last available visit; max = 7.57 years, mean = 2.81 years), and were classified as MCI converters according to previous definitions (32). All converter subjects had a Clinical Dementia Rating (CDR) score of 1. In contrast, those subjects who remained stable at follow-ups were classified as non-MCI converters. Sample processing followed standardized procedures (33). Briefly, plasma centrifuged from blood, as well as CSF were stored at −80 °C for later use. Core AD CSF biomarkers (Aβ40, Aβ42, t-tau, and p-tau181) were quantified using validated immunoassay platforms, with predefined thresholds used for AT(N) classification according to the 2018 NIA-AA Research Framework (34). For details on sample acquisition and collection of additional information, refer to previous publications (35–38).

### 2.2 Protein analysis in CSF and plasma using the SOMAscan panel

Paired CSF and plasma proteomic data from 1,369 ACE cohort participants were generated using the SOMAscan 7K Assay Kit in the context of the HARPONE project and the Global Neurodegeneration Proteomics Consortium (36–38). Briefly, this platform measures approximately 7,000 proteins per sample. This assay uses 50 μL of either CSF or plasma and employs modified DNA aptamers (SOMAmers) to quantify specific protein levels. Initially, proteins captured by immobilized aptamers, facilitated by streptavidin beads, and tagged with fluorescent markers. After removal of unbound proteins, the streptavidin beads are released using ultraviolet light. The protein-aptamer complexes were re-captured by monomeric avidin. The detail of the assay is described elsewhere (39). Data was reported in normalized relative fluorescence units (RFU) values using the adaptive normalization by maximum likelihood (ANML) approach, following previously described procedures (39). For the LTF gene (UniProt ID: P02788), two SOMAmers - Seq.14755.4 and Seq.2780.35 - were designed on the SOMAscan platform based on the target coding information provided by SomaLogic. These two SOMAmers have different dilution factors: 20% for Seq.14755.4 and 0.005% for Seq.2780.35 (**Supplement Table S1**). For clarity, we henceforth refer to Seq.14755.4 as LTF1 and Seq.2780.35 as LTF2 throughout this manuscript.

### 2.2. Statistical methods

#### a. Identification of the association between LTF expression and dementia diagnosis

Prior to statistical analysis, SOMAmer measurements were log-transformed and standardized using z-score normalization due to the long tail of the SOMAmer signal among participants. The normality of LTF1/2 distribution was evaluated with Shapiro.test function in R. To examine the association between LTF levels and AD-related biomarkers, the Spearman test was applied to compare the distributions of LTF SOMAmers, given their non-normal distribution. For LTF1, participants were further categorized into high (LTF1-high) and low (LTF1-low) groups to evaluate group-wise differences. Spearman correlation analysis was used to assess the relationships between LTF levels (in CSF and plasma), AD CSF biomarkers expression, and AD risk loci identified in a previous genome-wide association study (40). To address potential inaccuracies in p-values arising from tied ranks in Spearman correlation tests, genotype dosages were transformed into categorical values (0, 1, or 2), followed by Fisher’s exact test using the fisher.test function with simulate.p.value = TRUE and B = 10,000. All statistical analyses were performed using R (v4.3). To evaluated whether the confounding factor affects the association between LTF1 and AD biomarkers (Aβ42, p-tau181), linear models were fit with sex, age, age2, principal components and reference genes GAGE2A (seq.18268.5) or OPCML (seq.15622.13) signal as covariates. To perform principal components analysis, we filter out samples with more than 10% protein that have outlier abundance (absolute value ≥ mean ± 3 SD), the outlier value left are imputed using function imputPCA in R package missMDA (v: 1.20). PCA was calculated using function prcomp in R package stats (4.5.1).

#### b. Assessment of associations between clinical traits and LTF protein expression

Patients were categorized into case and control groups based on AT(N) biomarker classification (status). Individuals with an A-T-(N)- profile were assigned to the control group, while those with A+ (regardless of T or N status) were classified as cases. For further comparison, we also compared A+ v.s. A-, A+T+ v.s. A-T-, A+T+ v.s. A-T- (include A-T-(N)-). Participants were classified into three diagnostic categories: cognitively normal controls (including subjective cognitive decline (SCD)), mild cognitive impairment (MCI), and dementia. Due to the non-linear relation between LTF and CSF biomarkers, we categorize the samples into high and low according to the signals of LTF1 (High: > mean + SD; Low: < mean + SD) for correlation analysis. For LTF2, the outliers were Winsorized (mean ± 3 standard deviation).

#### c. Identification of the LTF central nervous system and systemic co-regulatory gene networks

To compare the LTF-associated genes with known co-regulated expression profiles, gene clusters containing 237 brain-expressed genes and 154 bone marrow–expressed genes were retrieved from the Human Protein Atlas database (41). LTF-associated proteins were identified using Spearman correlation analysis via the cor.test function in R. Genes with an absolute Spearman correlation coefficient greater than 0.1 and ranked among the top 250 proteins by correlation were selected for the positively (+) and negatively (-) correlated groups, respectively. Multiple testing correction was applied using the false discovery rate (FDR) method. Fisher’s exact test was used to assess enrichment by comparing the identified genes from the proteomic data with gene sets from the Human Protein Atlas database.

#### d. Cox proportional hazards models

We applied a series of Cox proportional hazards regression models to assess the association between LTF1/LTF2 and the risk of conversion to dementia. Three hierarchical models were constructed to evaluate the robustness of the associations under different levels of covariate adjustment: Model 1 (unadjusted model): LTF1 or LTF2 signal (LTF) was entered independently to estimate its association with dementia conversion. Model 2 (demographic-adjusted model): Each variable was included in a Cox model adjusted for age at lumbar puncture (Age_LP), sex, years of education (Education), and baseline Mini-Mental State Examination (MMSE) score. Model 3 (Fully adjusted model): In addition to the covariates in Model 2, we further adjusted for polygenic risk score (PRS), CSF phosphorylated tau (p-tau181) levels, and APOE genotype. Statistical tests were performed between the LTF1-high and -low groups. For LTF2, statistical significance was assessed by comparing samples with abundance above versus below the median. Hazard ratios (HRs), 95% confidence intervals (CIs), and p-values were reported for each model. Survival time was defined as the interval from MCI to dementia conversion or last follow-up assessment.

## 3. Results

### 3.1 LTF abundance in CSF and Plasma

To investigate whether LTF changes contribute to AD progression and the possibility to serve as a potential biofluid biomarker for AD, we accessed the plasma and CSF profiles of 1,369 individuals from the ACE CSF cohort. In this cohort, two samples lacked detectable signals for both LTF-targeting SOMAmers. Analysis included a total of 792 females and 577 males. The subjects consisted of 113 (8.27%) healthy controls; 833 (60.9%) diagnosed with MCI at baseline–of whom 44.1% converted to dementia during the follow-up visit, with CSF samples collected within 3 months of first visit–and 394 (28.2%) subjects diagnosed with dementia at the time of the lumbar puncture (**Supplement Table S2**). The mean age at the time of lumbar puncture was 72.7 ± 8.8 years. As expected, the frequency of the *APOE* ε4 allele was higher in the dementia group (21.9%) compared to MCI group (19.9%) and control group (13.1%). Detailed demographic and clinical characteristics are summarized in **Supplement Table S2**. We first investigated the distribution of all SOMAmers and noticed that distribution patterns of most SOMAmers did not follow a normal distribution including LTF1/2 in both CSF and plasma (**Fig. 1A**). For each individual, two SOMAmers that target LTF were measured in both CSF and plasma. For LTF1 (seq.14755.4) most individuals exhibited low expression in both CSF (132 ± 79) and plasma (370 ± 469) , while a small subset showed high expression in either fluid (**Fig. 1A, B**). The other SOMAmer (LTF2; seq.2780.35) displayed high expression in plasma (1,260 ± 4,703) for a small group of individuals (**Fig. 1A, C)** and another group of samples showed CSF-specifically high expression (CSF: 19,951 ± 20,804). This result indicates that LTF1 and LTF2 possibly represent two different isoforms or different conformational/binding status of LTF. To further confirm this hypothesis, we compared LTF expression between the two SOMAmers. The correlation analysis reveals the signal between the two SOMAmer shows weak significant correlation only in plasma (Spearman’s ρ = 0.22; p < 0.001), while there was almost no significant correlation in CSF (Spearman’s ρ = 0.01; p > 0.05; **Fig 1D**). Meanwhile, results revealed no significant correlation between CSF and plasma (Spearman’s ρ = 0.02; p > 0.05) for LTF2, with a similarly negligible negative correlation for LTF1 (Spearman’s ρ = -0.09; p < 0.01; **Fig 1D**). To examine whether the distribution of LTF1 in CSF is associated with blood-brain barrier integrity, we performed Spearman correlation analysis between LTF1, LTF2 and the albumin quotient (Qalb; a measure of blood–brain barrier (BBB) leakage), revealing that only CSF LTF1 levels (Spearman’s ρ = 0.1; p < 0.001; **Fig. 1E**), not LTF2 (Spearman’s ρ = -0.01; p > 0.05), weakly associated with Qalb.

**Fig. 1.**
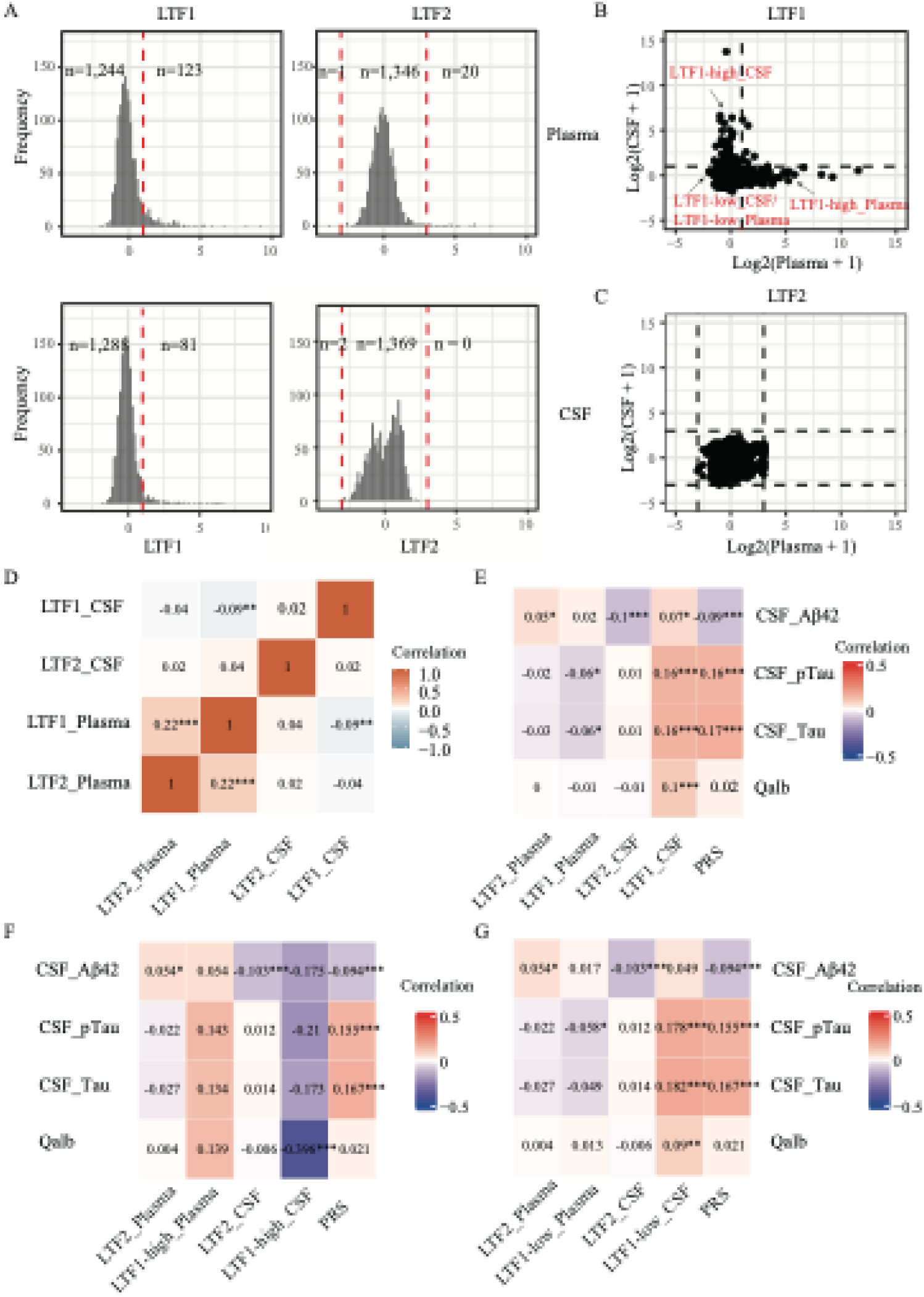
Distributions of SOMAmer signals targeting lactoferrin (LTF) and their associations with Alzheimer’s disease (AD) biomarkers. **A**: Distributions of two SOMAmers targeting LTF (LTF1 on the left, LTF2 on the right) in plasma (top) and CSF (bottom). Red dotted lines indicate the cutoffs defined as mean + standard deviations for LTF1 and mean ± 3 standard deviations for LTF2. All p-value were calculated by Shapiro.test for normality test. **B–C**: Comparisons of LTF1 and LTF2 signals between CSF and plasma; dotted lines represent the cutoffs shown in panel A. **D**: Heatmap showing the Spearman correlation between LTF1 and LTF2. **E–G**: Heatmaps showing Spearman correlations between the two SOMAmers, polygenic risk scores (PRS), and AD biomarkers. Panel E shows results for all samples; panels F and G show correlations after stratifying by LTF1 level into high and low for LTF1, respectively. *, p < 0.05; **, p < 0.01; ***, p < 0.001.

### 3.2 LTF Levels and AD Biomarkers

We sought to determine whether LTF expression is associated with established CSF AT(N) biomarkers in the ACE CSF cohort. To address this, we performed Spearman correlation analysis between LTF1 and LTF2 signals with Aβ42, p-tau181 and t-181 (**Fig 1E**). We found that LTF1 in CSF showed a weak positive correlation with total-tau (Spearman’s ρ = 0.16; p < 0.001), p-tau181 (Spearman’s ρ = 0.16; p < 0.001) and a negligible correlation with Aβ42 (Spearman’s ρ = 0.07; p < 0.05). In contrast, LTF2 showed a weak negative correlation with Aβ42 (Spearman’s ρ = -0.10; p < 0.001) and no significant association with p-tau181 (Spearman’s ρ = 0.01; p > 0.05) or t-tau (Spearman’s ρ = 0.01; p > 0.05). Based on the distribution patterns of LTF1 and LTF2 in CSF and plasma (**Fig. 1A**), we hypothesized that the weak correlation might reflect the heterogeneity in expression across subgroups of participants. LTF1 displayed a non-normal distribution, suggesting it may be expressed in only a subset of individuals. LTF2 appears to follow a normal distribution, with a few distinct outliers. To test this hypothesis, we stratified the samples based on LTF1 signal intensity: high (≥ mean + SD) and low (< mean + SD) and remove the outliers for LTF2. We then repeated the correlation analysis. For LTF2, results remained unchanged (**Fig 1F**; **Fig S1**). In the LTF1-low group, the associations with AD biomarkers were stable (**Fig 1G**; **Fig S1**), however, for LTF1-high groups, the direction of the associations reversed, and correlation coefficient increased, particularly for Aβ (Spearman’s ρ = -0.175), and p-tau181 (Spearman’s ρ = -0.21, **Fig 1F**; **Fig S1**). Notably, these coefficients were slightly stronger than those observed between AD polygenic risk score and AD biomarkers (variance explained increase by ∼2.0% for p- tau181, 0.2% for t-tau, and 2.2% for Aβ42). Our recent work showed that variability in CSF proteomes is dominated by a ventricular-volume–related dilution axis, implying that CSF biomarker signals can be confounded by physiological variability (42). To investigate its impact on the LTF–AD-biomarker relationship, we first examined correlations of LTF1/LTF2 with the first two proteomic principal components (PC1–PC2) and with reference genes (GAGE2A, OPCML). The result reveals LTF1 correlated with PC1 in the low-expression group (Spearman’s ρ = −0.373, p < 0.001; **Fig. S2**) but not in the high-expression group (ρ = 0.009, p > 0.05; Fig. S2). LTF2 showed no association with PC1 (Spearman’s ρ = −0.004, p > 0.05; **Fig. S2**)

To assess the influence of potential confounders, we re-estimated the associations between CSF LTF1/LTF2 and AD biomarkers using linear models that adjusted for sex, age, age², proteomic principal components (PC1–PC2), and reference-gene abundance (**Supplementary Tables S3–S6**). After adjusting for PC1, LTF1 was not associated with Aβ42 (β = −0.005; p = 0.089; **Supplementary Table S3**), but it remained negatively associated with p-tau (β = −0.167; p < 0.001; **Supplementary Table S3**). In stratified analyses, LTF1 continued to associate with p-tau in the high-expression group after PC1 adjustment (β = −0.144; p = 0.003), whereas the apparent positive association in the low-expression stratum (β = 0.383; p < 0.001) disappeared after additional adjustment for reference genes (β = 0.010; p = 0.837) and show negative association after adjustment for PC1 (β = -0.167; p = 0.001). For LTF2, the association with Aβ42 (β = −0.112; p < 0.001) was partially attenuated by reference-gene adjustment (β = −0.087; P < 0.001) but was unchanged by PC1 (β = −0.113; p < 0.001) (**Supplementary Table S6**). Collectively, these results suggest that the LTF1 signal in the low-expression group largely biased by physiological (dilution) variation before adjustment, whereas variation in high-expression LTF1 and in LTF2 is more likely to be pathologically relevant.

We then compared the LTF1 and LTF2 signal levels across participants with different pathological features. The results showed that LTF2 levels in CSF were significantly associated with AD status defined by AT(N) biomarker (p = 0.0051; **Fig S3**) and show trend toward an association with clinical symptom syndromic state (Kruskal-Wallis, p = 0.050; **Fig S3**), while LTF1 levels were not. To determine the biological relevance of these SOMAmer signals, we performed pathway enrichment analysis on the top co-regulated proteins for LTF1 and LTF2. For LTF1 (in both plasma and CSF), enriched terms could not be found before stratification, however, in the LTF1-high group, we observed enrichment for neural-related pathways in the plasma analysis, including “neurogenesis”, “generation of neurons”, “neuron projection development”, and “neuron differentiation” (**Fig 2A**). Proteins correlated with LTF1 in CSF were enriched in immune-related categories such as “blood microparticle” and “complement and coagulation cascades” (**Fig 2C**). To further explore the differences of high and low populations, we analyzed the LTF-associated protein networks using SomaScan data. Correlation analyses between LTF1/LTF2 and all other quantified proteins were performed, and significantly coregulated proteins were compared with LTF interactors identified in single-cell RNA-seq data from brain and bone marrow. Before stratification, no enrichment overlap was found for LTF1 in CSF or plasma. However, in the positive group, significant overlap was observed with bone marrow–derived co-regulated genes (**Fig 3**).

**Fig. 2.**
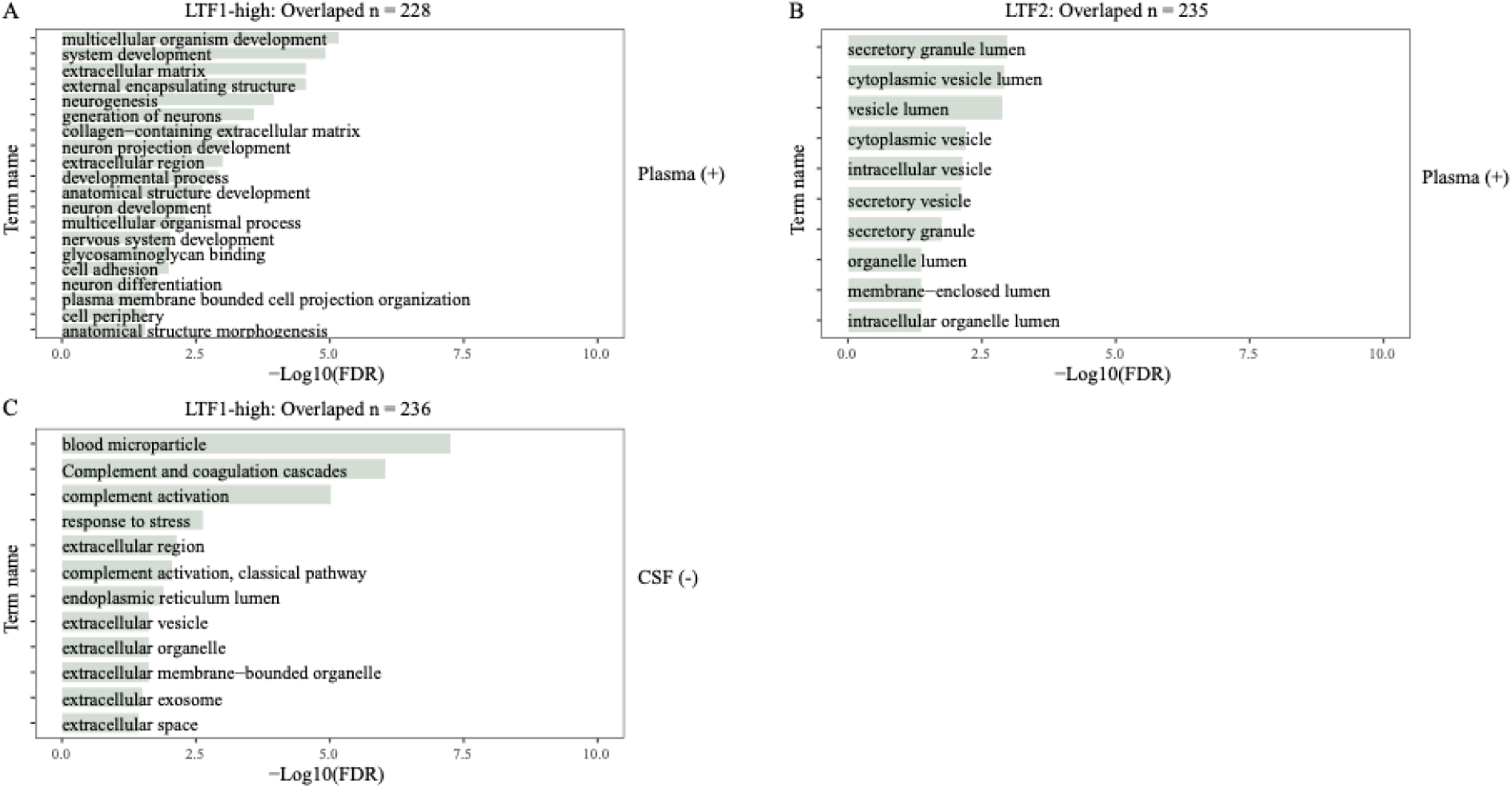
GO enrichment analysis of lactoferrin-associated proteins identified from SOMAscan plasma and CSF proteomic data. **A:** Significantly enriched Gene Ontology (GO) biological process terms among proteins positively correlated with the plasma-derived LTF1 signal in individuals classified as + (n = 226). **B:** GO cellular component terms enriched among proteins positively correlated with the plasma-derived LTF2 signal (n = 226). **C:** GO biological process and cellular component terms enriched among proteins negatively correlated with the CSF-derived LTF1 signal in the LTF1-positive group (n = 236). Enrichment significance is represented by -log10(FDR)-adjusted p-values.

**Fig. 3.**
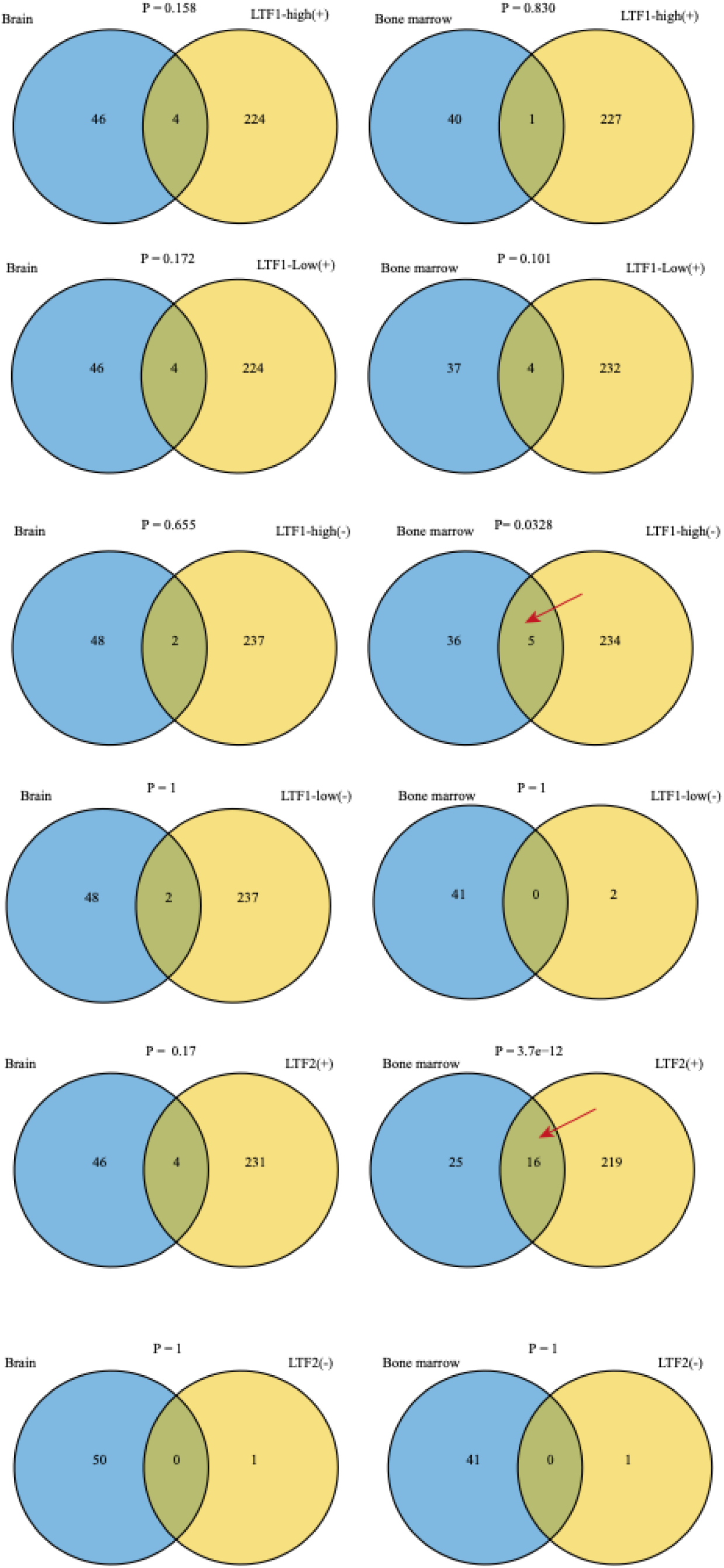
Overlap between LTF-derived correlation signatures in plasma and tissue-specific co-expression modules. Columns show tissue modules from the Human Protein Atlas database (Brain, #237 and Bone marrow, #154 blue circles). Rows stratify our signatures by (i) LTF1 expression stratum—LTF1-high vs LTF1-low and (ii) correlation sign with the LTF signal: “+” = top positively correlated genes, “−” = top negatively correlated genes. For LTF2, correlations were computed across all samples (no expression stratification). In each panel, the right circle is the LTF-derived signature (yellow) and the left circle is the tissue co-expression set (blue). Statistical significance was assessed with Fisher’s exact test (background = all tested genes after QC, n = 5,419).

LTF2 showed modest enrichment for terms like “secretory granule lumen” and “vesicle lumen” (**Fig. 2B**). Overlap with bone marrow-derived networks was also confirmed. Together, these findings suggest that LTF1 and LTF2 are associated with distinct proteomic profiles and biological processes, possibly reflecting different aspects of the LTF locus biology and AD pathology. Importantly, the diagnostic relevance of LTF1 appears to be limited to specific subgroups of samples that have higher expression for LTF1 signal.

### 3.3 CSF and Plasma LTF Levels Are Not Associated with AD Progression or Genetic Risk

We next examined whether LTF expression is associated with the progression from mild cognitive impairment (MCI) to dementia. Using Cox regression models, we found that neither LTF1 nor LTF2 expression in cerebrospinal fluid (CSF) or plasma was significantly associated with disease progression. To account for potential confounding variables, we implemented nested Cox regression models. The second model included adjustments for age, sex, years of education, and MMSE score, while the third model further adjusted for polygenic risk score (PRS), p-tau181 levels, and APOE genotype. Across all models, LTF1 and LTF2 levels in both CSF and plasma were not significantly correlated with conversion from MCI to dementia (**Fig. 4**).

**Fig. 4.**
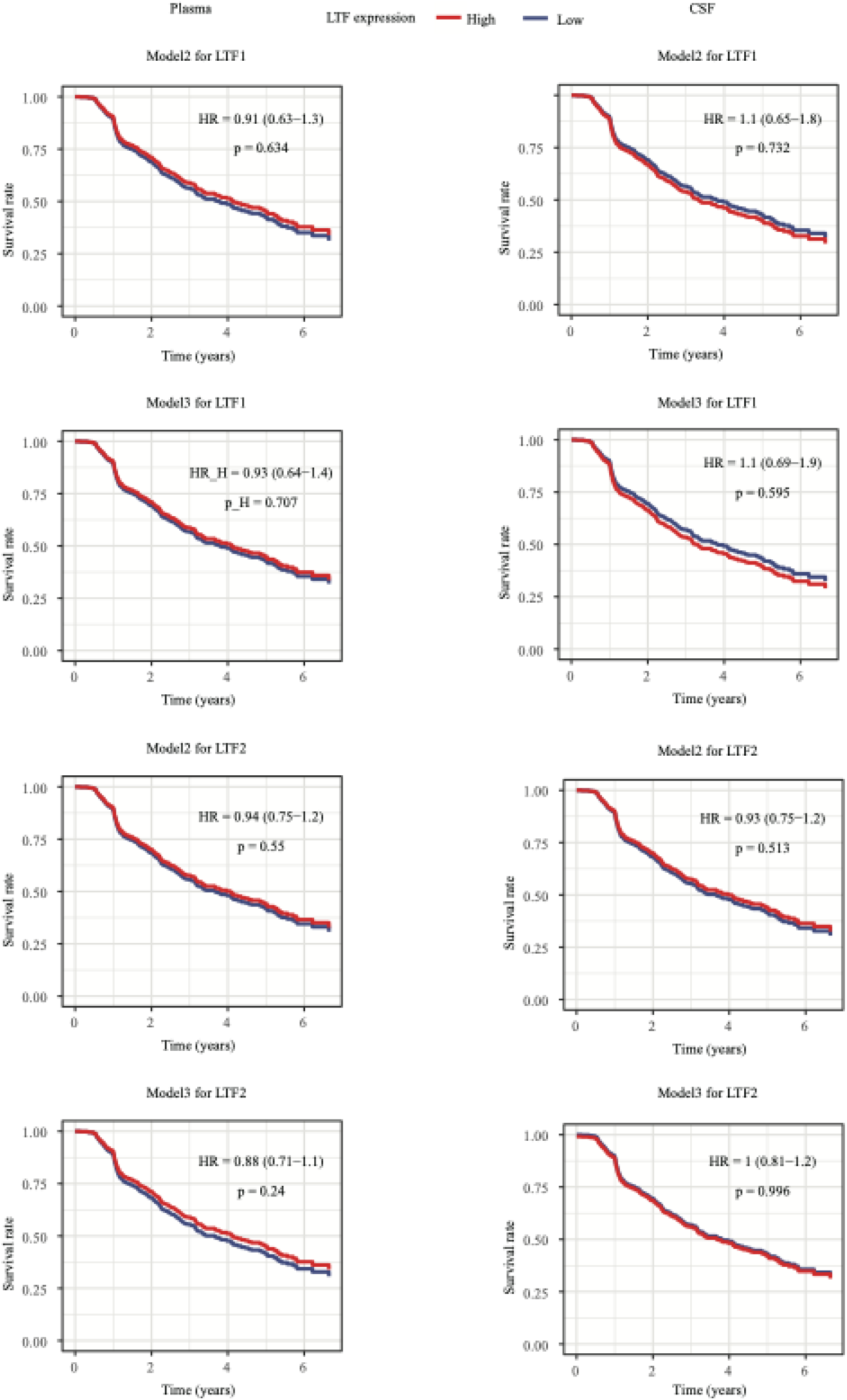
Survival analysis for lactoferrin (LTF) using nested models. The left column shows results based on plasma data; the right column shows results based on CSF data. For LTF1, survival rates were compared between LTF1 high and LTF1 low. For LTF2, samples were split by the median (High: ≥ median; Low: < median). Model1: Surv(Time(years), status) ∼ LTF + Age_LP + sex + education + mmse_csf; Model2: Surv(Time(years), status) ∼ LTF + csf_ p_tau + PRS + APOE + Age_LP + sex + education + mmse_csf. HR represents the hazard ratio of the positive group, 95% confidence intervals is included, and p is the corresponding p-value.

Additionally, we assessed whether variation in LTF expression could be driven by genetic risk for AD. However, no significant associations were found between AD-associated genetic variants and LTF expression levels in either CSF or plasma. These findings suggest that, despite the known roles of LTF in immune response and neuroinflammation, LTF expression levels in CSF and plasma do not appear to reflect AD progression or genetic susceptibility. This may indicate that LTF biomarker potential in CSF and plasma may be limited to identifying the presence of disease pathology, rather than providing a biomarker read for disease risk phenoconversion or genetic risk stratification in AD.

## 4. Discussion

Salivary LTF levels remain a controversial candidate biomarker for AD. To further investigate the potential role of LTF as a biomarker for AD, we examined LTF expression in both systemic and central compartments in the context of the AD diagnostic continuum in the largest dataset reported to date (N= 1,367). Our rationale was that if salivary LTF levels are directly associated with AD, similar associations might be detectable in other biological fluids – particularly in CSF, which is in closer contact with the neuropathological changes characteristic of the disease.

Our study was inspired by previous work suggesting a potential link between LTF and AD. Specifically, a 2017 study conducted a cross-sectional analysis using saliva samples and proposed salivary LTF as a promising biomarker to differentiate mild cognitive impairment (MCI) and AD from healthy controls (27). Two subsequent studies by the same group further supported this finding: one demonstrating that salivary LFT could distinguish AD from other forms of dementia (28), and the other showed that LTF abundance is closely related to AD pathology (29). Additional support came from a pilot study that reported significant associations between salivary LTF levels and cognitive performance, as measured by the Digit Span Memory Test and Mental Rotation Test scores (43). Collectively, these studies suggest that reduced LTF levels in saliva may serve as a non-invasive biomarker for AD diagnosis. However, a study from an independent laboratory failed to replicate these findings in a consecutive, mixed memory clinic cohort (30), casting doubt on the reliability of salivary LTF as an AD biomarker. Each of these investigations were based on relatively small cohorts (N= 17-274)(27–30, 43), which may reduce statistical power and contribute to inconsistent findings (44). Thus, it remains to be determined whether changes in LTF levels are part of the causal chain driving neurodegeneration, or merely epiphenomena reflecting systemic or local inflammation. Additionally, the function of LTF is complex – it can exhibit both pro-inflammatory and anti-inflammatory properties depending on the context of response (31).

Despite growing interest in LTF as a potential biomarker or mediator in AD, several limitations complicate its study across different biological compartments. Beyond its role in inflammation, LTF can act as a mediator of both innate and adaptive responses by promoting the maturation, differentiation and activation of T- B-lymphocytes (45). Its molecular complexity further challenges interpretation: LTF exists in multiple isoforms α, β, γ, and δ-LTF. The canonical isoform is primarily expressed in neutrophils and epithelial cell, secreted into exocrine inflamed tissue and fluid, while δ-LTF localizes to cytoplasm and act as a transcription factor (46, 47). Additionally, Importantly, iron-binding status further complicates LTF biology. LTF exists in different iron-binding states including the iron-free (apo-LTF), native and iron-saturated forms (holo-LTF), which may have distinct biological functions and activities (48, 49). Apo-LTF mainly sequesters iron, while holo- LTF can activate immune signaling pathways via receptors such as LRP1 and CXCR4, influencing MAPK, AKT, and NF-κB pathways (50, 51). Proteolytic cleavage of LTF can also generate biologically active antimicrobial peptides known as lactoferricins, which exhibit different functional properties compared to native LTF (52). Despite this molecular complexity, most current studies-including those evaluating its potential as an AD biomarker - use ELISA kits based on a single LTF antibody, which typically quantifies total or a single protein isoform, without distinguishing among various functional forms. Moreover, these studies often rely on correlation analyses between LTF levels and disease biomarkers, without addressing the biological significance or mechanistic relevance of inter-individual variation in LTF expression.

In our study, we aimed to explore the relationship between LTF levels in CSF and plasma with AD status, disease progression and AT(N) biomarkers. Unlike previous work focusing on salivary LTF, we used data derived from high-throughput SOMAscan proteomics on larger, well-characterized cognitive and biomarker cohorts. Our findings suggest that the relationship between LTF abundance and AD biomarkers is complex and compartment specific.

In our hands, lactoferrin expression in CSF and Plasma shows a complex and inconsistent association with AD biomarkers. Three key observations highlight the complexity of interpreting LTF measurements using SOMAmer technology: 1. Lack of consistency between SOMAmers: The two SOMAmers targeting to LTF (LTF1 and LTF2) showed markedly different distributions and negligible correlation in both CSF and plasma (**Fig 1D**). 2. Weak systemic-central correlation: Correlation between LTF SOMAmer signals between CSF and plasma was weak (**Fig 1E**). 3. Inconsistent association with AD biomarkers: In CSF, LTF1 displayed weak and even opposing correlations with AD biomarkers p-tau depending on LTF-high vs. low subgroups (**Fig 1F**, **G**) which may be caused by the variability in CSF protein abundance. Meanwhile, LTF1 is correlated with Qalb, but not for LTF2, suggesting that LTF1 may be more sensitive to barrier/inflammation-related signals, whereas LTF2 likely captures a different conformational or compartment-specific facet of LTF. Notably, CSF LTF2 levels were significantly higher in A+ than A-T-N- individuals (**Fig.S2**, p = 0.0019), and this difference was replicated in comparisons between A+T+ and A-T- groups (p = 0.0068). This pattern was not observed for LTF1. Together, these findings indicate that LTF measurements obtained with SOMAmer reagents are heterogeneous across targets and fluids.

The most prominent studies evaluating LTF as an AD biomarker were conducted using saliva samples. Carro, et al. (27) first reported positive associations between salivary LTF and CSF Aβ42, and negative association with t-tau, suggesting that lower levels of salivary LTF are associated with increased markers of AD pathology. Later studies from the same group supported the discriminatory power of salivary LTF between AD and other dementia types (28), and also linked it to cortical Aβ load and brain integrity in aging adults (29). A separate pilot study (43) found lower salivary LTF in participants with much more severe cognitive impairment. In contrast, our data from CSF and plasma cohorts challenge these findings, but do not necessarily negate them, as we were unable to replicate their work in saliva. We found there is higher CSF LTF2 levels in MCI dementia compared with control (Kruskal-Wallis test; p = 0.05). These findings were consistent with those of another independent study (30), which analyzed 222 matched saliva and CSF samples from a consecutive, mixed memory clinic population and found both CSF and saliva LTF expression have higher LTF expression in MCI and AD participants than healthy control although the difference is not statically significant. This discrepancy may arise from difference in sample origin, population heterogeneity, or the biological nature of LTF secretion in different compartments. It is possible that LTF expression in CSF do not reflect the association previously described for salivary LTF in AD. If the salivary LTF finding is genuine, it is most likely capturing a biological signal that is not adequately measured by SOMAscan assays in CSF or plasma. Moreover, the lack of saliva samples in our study limits direct comparability.

The molecular and multifunctional complexity of LTF likely contributes to these inconsistent results. LTF is involved in both innate and adaptive immune responses, influencing T- and B-cell activation (45). It exists in multiple isoforms (α, β, γ, δ), with distinct localization and functions. For example, δ-lactoferrin acts as a transcription factor and is localized in the cytoplasm (47). Meanwhile, iron-binding status of LTF also potentially impact on it biological functions and activities (48, 49). Another aspect is the technical considerations in SOMAmer-Based measurement. According to SomaLogic documentation, LTF1 targets amino acids 20 – 710, while LTF2 spans the full LTF sequence. These reagents may bind distinct epitopes, requiring different dilutions and potentially detecting different isoforms or modifications of LTF. Consistent with this notion, LTF1 and LTF2 show distinct associations with Qalb, suggesting that structural and conformational differences may underlie compartment-specific behavior and, consequently, their differential relationships with disease biomarkers. This distinction may partly explain the divergent results we observed.

Our study benefits from two notable key methodological strengths, multi-tissue protein profiling, enabling cross-compartment comparison; and large-scale proteomic-level analysis, providing consistently measured insights into disease relevant proteins.

However, several limitations must be acknowledged. First, our sample size, especially for healthy controls, may limit our statistical power. Second, though we had access to CSF and plasma proteomics, we lack this data in a cohort of saliva samples, which restricts our cross-compartment analysis and limits our interpretation of the data compared to previously published reports. We also acknowledge potential confounding introduced by unmeasured variables such as oral health, diet, or use of specific medications that may influence LTF levels or tissue secretion. Unfortunately, no isoform-level validation of SOMAmers exists to provide additional insight into the protein, protein-isoform, or component that is being measured, a critical consideration for proteins with known physical and functional diversity. Finally, we acknowledge that we have pulled our samples from a heterogeneous memory clinic population, which likely dilutes group-level effects that might otherwise be detectable.

Future work should include experimental validation of LTF isoforms and iron-binding forms across compartments, as well as mechanistic studies to clarify their roles in neuroinflammation and AD pathology.

Although lactoferrin shows some association with AD-related markers (particularly Aβ), its value as a biomarker remains uncertain due to its molecular complexity, compartment-specific behavior, and methodological limitations. Integrative multi-omics studies (36) emphasize the disconnection between protein abundance and genetic risk, reinforcing the need to explore regulatory and post-translational mechanisms. While exogenous LTF has shown therapeutic effects in preclinical models (53), such interventions may only benefit individuals with active LTF-related pathways. Identifying LTF-high subgroups in CSF or plasma could help stratify patients and tailor future treatments. However, it requires improved testing systems that could differentiate between LTF isoforms, their fragments, and their iron-binding states for disentangling the systemic and brain-specific roles of LTF in the pathophysiology of AD and related dementias. Ultimately, a deeper understanding of LTF’s systemic and brain-specific roles is crucial for developing effective, personalized strategies for AD diagnosis and therapy.

## Data Availability

All data produced in the present study are available upon reasonable request to the authors

## Contributors

Conceptualization: Agustin Ruiz, Valentina R. Garbarino; Formal analysis: Feiyang Zhao; Funding acquisition: Agustin Ruiz, Sudha Seshadri, Merce Boada, Jose E. Cavazos, Sergi Valero, Amanda Cano and Maria Victoria Fernandez; Methodology: Feiyang Zhao, Agustin Ruiz; Project administration: Feiyang Zhao, Agustin Ruiz; Supervision: Merce Boada, Jose E. Cavazos, Agustin Ruiz, Sudha Seshadri; Validation: Feiyang Zhao; Visualization, Feiyang Zhao; Writing – original draft: Feiyang Zhao, Agustin Ruiz, Yaxi Wang; Writing – review & editing: Valentina R. Garbarino, Tiffany Kautz, Agustin Ruiz, Sudha Seshadri, Jose E Cavazos, Yaxi Wang, Eva Beckett, Feiyang Zhao; Proteome database generation and curation, APOE genotyping GWAS analysis and PRS construction: Raquel Puerta, Pablo Gacia Gonzalez, Maria Victoria Fernandez and Amanda Cano; Clinical and molecular data acquisition: Merce Boada, Amanda Cano, Adelina Orellana, Pilar Sanz and Maria Victoria Fernandez. All authors had access to the data and accept the responsibility to submit for publication. All authors have read and approved the final version of the manuscript.

## Acknowledgments

We would like to thank the patients and controls who participated in this project. We are also grateful to the ACE Alzheimer Center Barcelona Memory Clinic, the clinical trial and nurse station staff involved in participant evaluations and case management, for their passionate dedication to the patients and families contributing to this research.

---

**Fig. S1.**
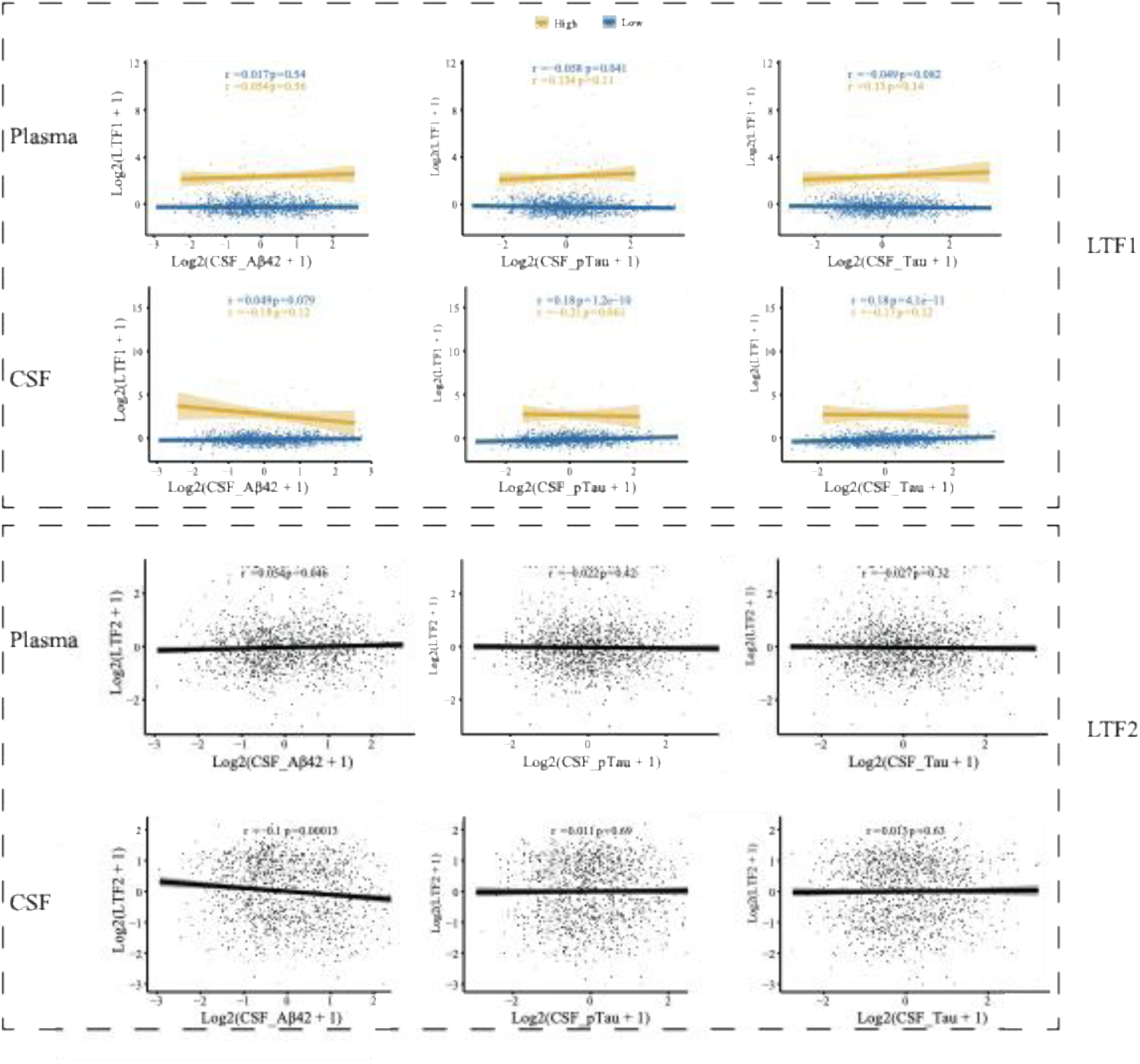
Scatter plots showing the correlations between LTF1/LTF2 and AD biomarkers (Aβ42, p-tau, and t-tau). The top row shows correlations with LTF1, and the bottom row with LTF2. LTF1 is stratified by LTF1 expression—LTF1-high (yellow) vs LTF1-low (blue). For LTF2, correlations were computed across all samples. Spearman correlation coefficients (r) and corresponding *p* values (*p*) are indicated in each panel.

**Fig. S2.**
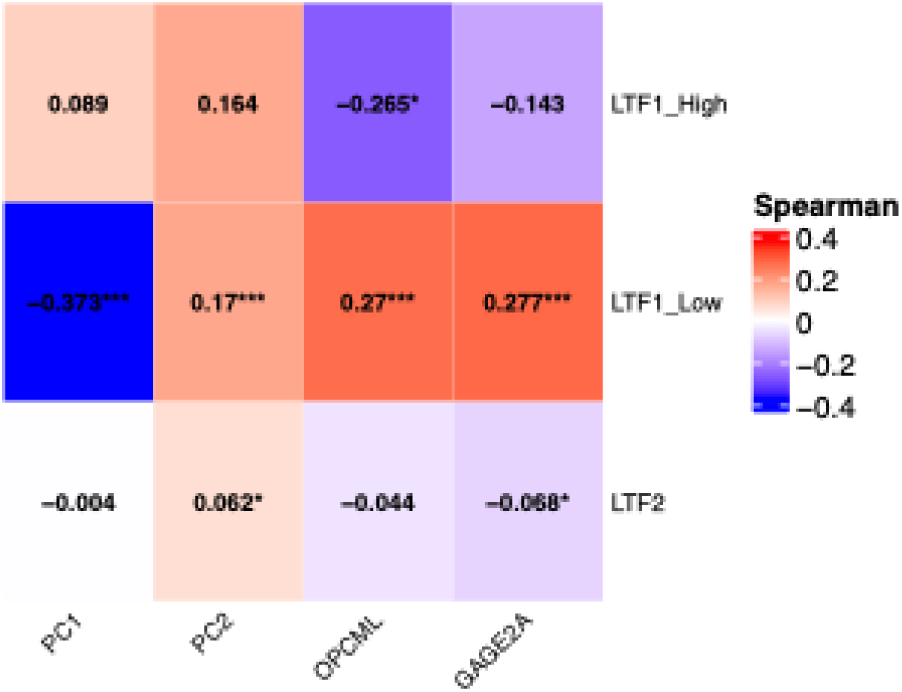
Spearman correlation heatmap between CSF LTF1-/LTF2 and the first two proteomic principal components (PC1–PC2) and reference genes (OPCML, GAGE2A). Cells display Spearman’s ρ; color encodes effect size. Significance: *, p < 0.05; **, p < 0.01; **, p < 0.001.

**Fig. S3.**
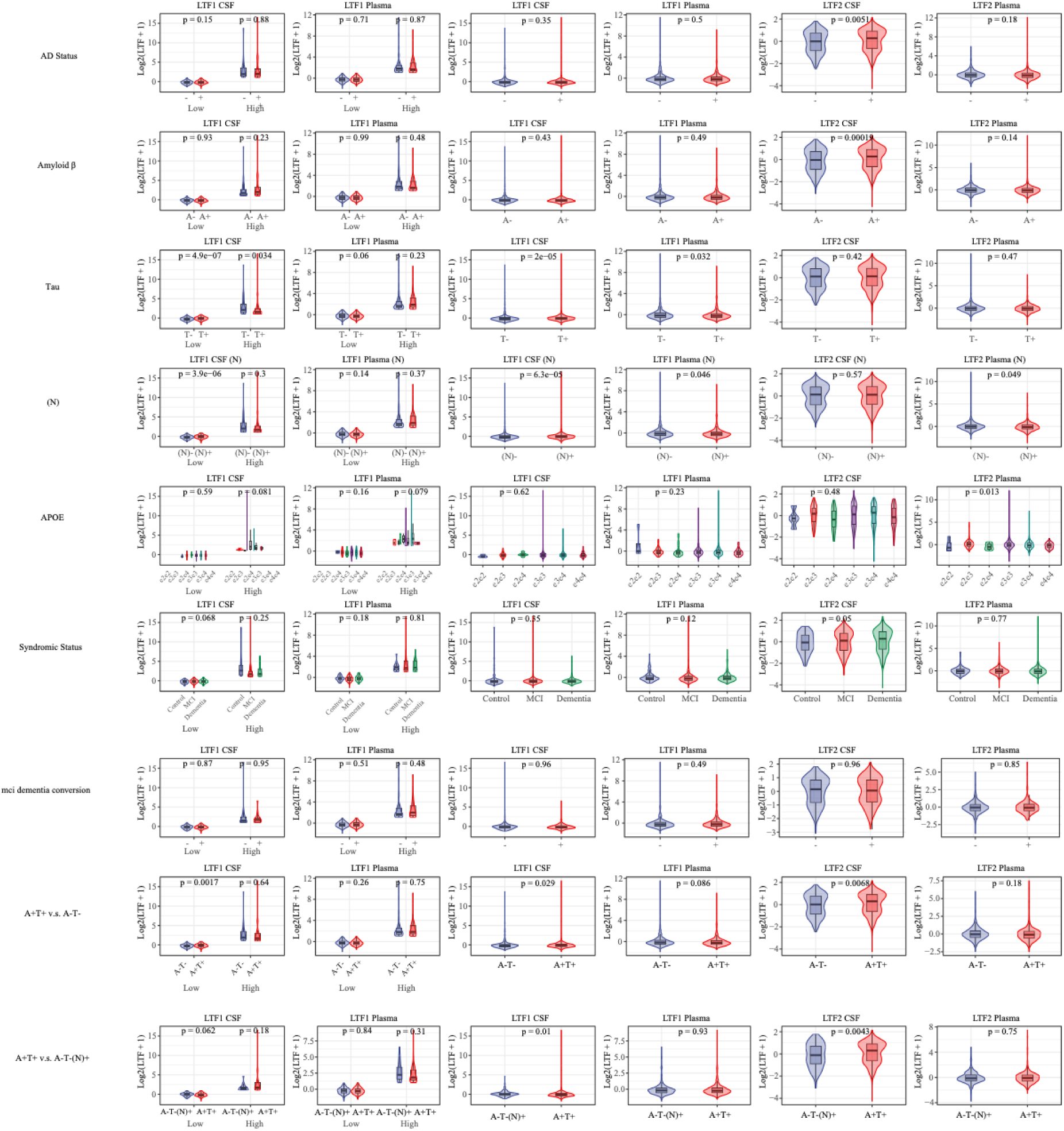
Comparison of LTF1 and LTF2 levels across AD biomarker profiles, clinical diagnosis, APOE genotypes, and longitudinal conversion status. Each row represents a comparison between groups stratified by APOE genotype, dementia diagnosis, A/T/N biomarker positivity, or longitudinal conversion status. Columns 1 – 2 show CSF and plasma LTF1 levels stratified by LTF1 abundance (high, low) respectively; columns 3 – 4 show the CSF and plasma LTF1 levels before stratification. Columns 5 – 6 show CSF and plasma LTF2 levels. In each plot, different color were used to discriminate the status of diagnosis, AD biomarkers, APOE genotypes or conversion status respectively. Statistical comparisons were performed using the Kruskal-Wallis test.

## Supplement Table

**Table S1.** Annotation of the two SOMAmers that target to LTF.

**Table S2.** Demographic and clinical data of the participants in the ACE cohort.

Abbreviations: LP: lumbar puncture; MCI, mild cognitive impairment; AD, Alzheimer’s disease dementia; MMSE, Mini Mental State Examination; LTF, Lactoferrin; NA, not applicable; SD, standard deviation.

**Table S3.** This table show the results of the linear regression between csf Aβ42, p-tau and LTF1 in CSF using sex, age, age^2^, reference gene, PC1, PC2. Results are shown for models adjusting for each covariate individually and for a fully adjusted model including all covariates simultaneously.

**Table S4.** This table show the results of the linear regression between csf Aβ42, p-tau and LTF1 in CSF using sex, age, age^2^, reference gene, PC1, PC2 as covariates in LTF1 high express population. Results are shown for models adjusting for each covariate individually and for a fully adjusted model including all covariates simultaneously.

**Table S5.** This table show the results of the linear regression between csf Aβ42, p-tau and LTF1 in CSF using sex, age, age^2^, reference gene, PC1, PC2 as covariates in LTF1 low express population. Results are shown for models adjusting for each covariate individually and for a fully adjusted model including all covariates simultaneously.

**Table S6.** This table show the results of the linear regression between csfA β42, p-tau181 and LTF2 in CSF using sex, age, age^2^, reference gene, PC1, PC2 as covariates. Results are shown for models adjusting for each covariate individually and for a fully adjusted model including all covariates simultaneously.

